# Comparison of the prevalence and associated factors of chronic kidney disease diagnosed by serum creatinine or cystatin C among young people living with HIV in Uganda

**DOI:** 10.1101/2024.09.02.24312932

**Authors:** Esther M Nasuuna, Laurie A Tomlinson, Robert Kalyesubula, Chido Dziva Chikwari, Barbara Castelnuovo, Yukari C Manabe, Damalie Nakanjako, Helen A. Weiss

## Abstract

**Introduction:** Young people living with HIV (YPLHIV) are at increased risk of developing chronic kidney disease (CKD) which is associated with high mortality and morbidity. Early diagnosis is important to halt progression. We aimed to estimate the prevalence and factors associated with CKD among YPLHIV in Kampala, Uganda, and to compare serum creatinine and cystatin C for early diagnosis of CKD in this population.

**Methods:** A cross-sectional study with YPLHIV aged 10 to 24 years was conducted in seven HIV clinics. Participants provided a urine and blood sample to measure urinary albumin, proteinuria, serum creatinine and cystatin C levels at baseline and after three months. The estimated glomerular filtration rate (eGFR) was calculated using CKDEPI 2021, Cockroft-Gault and bedside Schwartz equations using creatinine or cystatin C. The albumin creatinine ratio (ACR) and proteinuria were measured. CKD was defined as either eGFR <60ml/min/1.73m^2^ or <90ml/min/1.73m^2^ or ACR above 30mg/g on two separate occasions. Univariable and multivariable logistic regression were used to estimate adjusted odds ratios (aOR) and 95% confidence intervals (CI) for factors associated with CKD.

**Results:** A total of 500 participants were enrolled. Most were female (56%; n=280) and aged 10 to 17 years (66.9%; n=335). CKD prevalence ranged from 0-23% depending on the criteria, equation and biomarker used. Cystatin C-based equations estimated higher prevalence of CKD compared to creatinine-based ones. Prevalence of ACR above 30mg/g was 10.1% and of proteinuria 29%. Factors independently associated with CKD were age (aOR=1.42; 95% CI:1.30-1.51) and male sex (aOR=3.02; 95% CI:1.68-5.43).

**Conclusion:** CKD prevalence among YPLHIV varied substantially depending on definitions used and the current definition would likely lead to missed cases of CKD among YPLHIV. Estimating equations should be validated against measured GFR in YPLHIV and the optimal definition of CKD in this vulnerable population should be revised to optimise detection and opportunities for reducing disease progression.

## Introduction

Prevalence of chronic kidney disease (CKD) is increasing globally (1). CKD is defined as abnormalities in kidney structure or function present for three or more months (2). The Global Burden of Disease study estimates CKD prevalence at 9.1% (95% CI 8.5%-9.8%) with geographic variation (3). Studies in Sub-Saharan Africa (SSA) find prevalence ranging from 6%-48% depending on the population, the definitions used, and the measurements taken (4–6).

Young people living with HIV (YPLHIV) are at higher risk of CKD than young people not living with HIV (7). CKD risk is associated with high HIV viremia (>4000 copies per ml), severe immunosuppression (CD4 cell count <200 cells/ml), infection with hepatitis C virus, diabetes, hypertension, use of drugs that treat opportunistic infections, and toxicity due to anti-retroviral therapy (ART) from tenofovir disoproxil fumarate (TDF) and indinavir (6, 8–11). Further, YPLHIV in SSA are particularly vulnerable to developing CKD compared to adults living with HIV due to late HIV diagnosis and initiation on ART, poorer adherence to ART complicated by high viremia and low CD4 cell counts (12–14).

CKD is associated with high morbidity and mortality as diagnosis is usually delayed, often occurring after kidney failure due to its insidious onset (15). Kidney failure can only be treated with expensive kidney replacement therapies that are not readily available in low and middle-income countries (16). Early diagnosis is important to minimise risk of progression to kidney failure and cardiovascular events (17).

Diagnosis of CKD is based on the level of glomerular filtration rate (GFR) and markers of kidney damage such as protein excretion into the urine shown by proteinuria or albuminuria (18). GFR can either be measured directly (mGFR) or estimated (eGFR) with a specific biomarker and one of the estimating equations (19). Most commonly, serum creatinine and cystatin C estimating equations are used to estimate GFR (18, 20). Serum creatinine is widely available and relatively cheap (21) but has limitations as it is influenced by muscle mass, physical activity and general health status (22) as well as high analytic variability (21). Cystatin C is not affected by these conditions as it is produced by most nucleated cells and has uniform generation despite individual differences in people and situations (22–24). However, it is affected by conditions of high inflammation, corticosteroid use and thyroid disease (25). Cystatin C more accurately estimated measured GFR compared to creatinine (26) in a large cohort study done across Uganda, Malawi and South Africa that recommended the use of Cystatin C in African populations (4).

Although YPLHIV are at high risk of CKD, little is known about CKD prevalence, the best biomarker to diagnose CKD and factors associated with CKD in this vulnerable group. Therefore, we sought to study this among YPLHIV in Kampala, Uganda.

## Methods

### Study design and setting

This cross-sectional study was conducted in the HIV clinics of seven urban public health facilities from the 12^th^ of April 2023 to 31^st^ January 2024 in Kampala, Uganda. These offer comprehensive HIV care to children (aged below 18 years) and adults (aged 18 years and above).

### Study population and sampling

The study included YPLHIV aged 10-24 years with presumed perinatal HIV infection (defined as being diagnosed with HIV before 10 years of age with self-report of no sexual debut or blood transfusion prior to diagnosis). Pregnant YPLHIV were excluded. Systematic random sampling was used to identify potential participants from all YPLHIV enrolled in the seven HIV clinics from electronic medical records. They were ordered by age at diagnosis and every third person invited to join the study. A sample size of 500 was powered to detect a prevalence of CKD between 16% and 24%.

### Study procedures

Eligible participants were invited to the HIV clinic through a phone call where they were screened, consented, and enrolled. A trained study team member conducted an interview with the participant and completed a questionnaire to record demographic information, symptoms, risk factors and the relevant medical history. Anthropometric measurements (mid-upper arm circumference (MUAC), weight and height) were taken. Weight was assessed using a digital weighing scale, height using a stadiometer and blood pressure (BP) using a digital BP machine with a paediatric cuff for younger participants. Body composition monitoring was conducted using bioimpedance impedance spectroscopy (BIS) to measure body fat, muscle mass and visceral fat. Participants provided a spot urine sample (20 mls) as well as 8 ml of venous blood. Urine dipstick was done at the facility to determine proteinuria and other urinary abnormalities. The samples were stored in a cooler box before transfer to the study laboratory on the same day.

### Laboratory methods and testing

In the laboratory, serum creatinine, urinary albumin and cystatin C levels were determined. Those with an albumin creatinine ratio (ACR) >30mg/g or eGFR <60ml/min/1.73m^2^ at baseline were followed-up after three months to confirm the KDIGO guideline-recommended clinical diagnosis of CKD. Cystatin C was measured by particle-enhanced immunoturbidimetric assay on Roche Cobas C311 platform with Tina-quant Cystatin C Gen.2. Creatinine was measured using the enzymatic calorimetric method using an isotope dilution mass spectroscopy (IDMS) traceable standard reference material on the Cobas Integra 400 plus machine with Creatinine Plus Version 2 (CREP2), Roche Diagnostics. The urine albumin was quantified using the immunoturbidimetric assay on the Roche Cobas C311 platform using Tinaℒquant Albumin Gen2, (Roche Diagnostics). Prior to testing, the machines were calibrated according to manufacturer instructions.

Urinalysis by dipstick was done with AYDMED urinalysis Reagent Test Strips (Sungo Europe B.V Amsterdam) to determine presence of urobilinogen, bilirubin, ketones, blood, proteins, nitrites, leucocytes, glucose, specific gravity, pH, and ascorbic acid (27). **Diagnosis of CKD** was based on the kidney disease improving global outcomes (KDIGO) guidelines (28), i.e. 1) markers of kidney damage such as an albumin: creatinine ratio >30mg/g, or 2) eGFR <60ml/min/1.73m^2^, with these abnormalities confirmed with a repeat test after three months (29). To explore CKD definition in this cohort that included children and where chronic disease had affected pubertal development and mean body and muscle mass, we primarily used a range of GFR estimating equations and eGFR cut offs that reflected contemporary practice for adults and children and/or sought to adjust for body size. eGFRscr was estimated using the following creatinine-based equations: CKD Epidemiology collaboration (CKDEPI) 2021 (30), the Bedside Schwartz (31), and Cockroft-Gault. eGFRcystc was estimated using the following cystatin C-based equations: Schwartz cystatin C (32) and CKDEPI 2012 (33). For completeness prevalence was also estimated using other relevant equations (Full Age Spectrum, CKDEPI40 and Pierce U25), and in combination with ACR and proteinuria. Since a normal GFR is between 90-120 ml/min/1.73m^2^, we also considered a eGFR cut off below 90ml/min/1.73m^2^ which is considered stage 2 CKD as abnormal in such a young population (34).

### Data management and statistical analysis

Data were collected in REDCap and analysed with STATA statistical software version 18 (STATA Corp USA). Viral suppression was considered as an HIV viral load below 1000 copies/ml. Hypertension was classified according to the AAP guidelines as being above the 95^th^ percentile for age and sex below 13 years and above 130/80 in those above 13 years (35). Muscle mass was abnormal if below 33.3 for males and 24.3 for females. Social economic status was divided into three using principal component analysis. Demographic data were summarised in percentages or means (standard deviation) and median (interquartile range). The distribution of eGFRs estimated with different equations was shown in a Kernel density plot. CKD prevalence diagnosed by either creatinine or cystatin C was calculated. Univariable logistic regression was used to estimate odds ratios (OR) of factors associated with CKD for each of the five equations used, respectively. All variables with p<0.2 in the univariable model, and a-priori identified variables known to be associated with CKD (age, sex, HIV viral suppression, blood pressure) were then included in a multivariable logistic regression model for each of the five equations.

### Ethical considerations

Ethical approval was received from the Uganda Virus Research Institute (UVRI) Research Ethics Committee (reference number GC/127/946), the Uganda National Council of Science and Technology (HS2578ES) and the London School of Hygiene and Tropical Medicine institutional review board (28797). Information about the study appropriate for adults, semi-literate adults and children was provided in an information booklet that was read to the participants and caregivers. All the participants more than 18 years of age provided a written informed consent. Those below 18 years of age provided assent and their caregivers provided written informed consent. If a child refused to provide assent even after their caregiver had provided consent, that child was not enrolled into the study. All participants had the option to withdraw at any point during the research. All participants with suspected CKD were referred to a nephrologist for management.

## Results

Of 532 YPLHIV invited to participate, 500 were enrolled as the 32 declined to participate (**Table 1**). The majority were female (56.0%; n=280), children aged 10-17 years (66.9%; n=335) and living in Kampala (58.9%; n=295). Females had better nutritional indicators than males - they were less likely to be underweight (26.4% vs 48.9%; p<0.001), not stunted (85.6% vs 76.9%; p=0.03), and to have normal mid upper circumference (92.9% vs 87.7%; p=0.05).

**Table 1:** Demographic characteristics of the study participants by sex.

|  | Male<br>N=220 (44%) | Female<br>N=280 (56%) | Total<br>N=500 |
| --- | --- | --- | --- |
| **Age (mean, SD)** | 16.5 (3.8) | 16.3 (3.5) | 16.4 (3.6) |
| **Age category** |  |  |  |
| Children | 143 (65.0) | 191 (68.2) | 334 (66.8) |
| Adults | 77 (35.0) | 89 (31.8) | 166 (33.2) |
| **Address** |  |  |  |
| Kampala | 123 (55.9) | 171 (61.1) | 294 (58.8) |
| Wakiso | 80 (36.4) | 99 (35.4) | 179 (35.8) |
| Other districts | 17 (7.7) | 10 (3.6) | 27 (5.4) |
| **Religion** |  |  |  |
| Christian | 158 (71.8) | 204 (72.9) | 362 (72.4) |
| Moslem | 62 (28.2) | 73 (26.1) | 135 (27.0) |
| Other | 0 (0.0) | 3 (1.1) | 3 (0.6) |
| **Social Economic Status** |  |  |  |
| Lowest | 78 (35.5) | 100 (35.7) | 178 (35.6) |
| Middle | 61 (27.7) | 94 (33.6) | 155 (31.00) |
| Highest | 81 (36.8) | 86 (30.7) | 167 (33.4) |
| **School going** |  |  |  |
| No | 58 (26.4) | 67 (23.9) | 125 (25.0) |
| Yes | 162 (73.6) | 213 (76.1) | 375 (75.0) |
| **Marital Status** |  |  |  |
| Married | 3 (1.4) | 16 (5.7) | 19 (3.80) |
| Never married | 217 (98.6) | 264 (94.3) | 481 (96.2) |
| **Tribe** |  |  |  |
| Ganda | 149 (67.7) | 174 (62.1) | 323 (64.6) |
| Other tribes | 65 (29.5) | 86 (30.7) | 151 (30.2) |
| Non-Ugandan | 6 (2.7) | 20 (7.1) | 26 (5.2) |
| **Weight mean (SD)** | 48.0 (12.4) | 49.4 (12.4) | 48.8 (12.4) |
| **Body Mass Index<sup>1</sup>** |  |  |  |
| Normal | 173 (79.0) | 214 (76.4) | 387 (77.6) |
| Underweight (<18.5kg/m<sup>2</sup>) | 38 (17.3) | 24 (8.6) | 62 (12.4) |
| Overweight (>25kg/m<sup>2</sup>) | 8 (3.7) | 42 (15.0) | 50 (10.0) |
| **Stunting\*\*** |  |  |  |
| Not stunted | 123 (76.9) | 184 (85.6) | 307 (81.9) |
| Stunted | 37 (23.1) | 31 (14.4) | 68 (18.3) |
| **Mid Upper Arm Circumference <sup>2</sup>** |  |  |  |
| Normal | 192 (87.7) | 260 (92.9) | 452 (90.6) |
| Malnourished | 27 (12.3) | 20 (7.1) | 47 (9.4) |
| **On TDF regimen** |  |  |  |
| No | 61 (27.7) | 66 (23.6) | 127 (25.4) |
| Yes | 159 (72.3) | 214 (76.4) | 373 (74.6) |
| **Virally suppressed** |  |  |  |
| Yes | 195 (89.5) | 247 (88.5) | 442 (88.9) |
| No | 23 (10.5) | 32 (11.5) | 55 (11.1) |
| **Muscle mass <sup>2</sup>** |  |  |  |
| Normal muscle mass | 164 (82.0) | 235 (87.7) | 399 (85.3) |
| Abnormal muscle mass | 36 (18.0) | 33 (12.3) | 69 (14.7) |

\* Living outside Kampala/Wakiso region. # included those with no religion and those of African traditional religion.
\*\*Only those aged less than 19 years. 1 one missing, 2. 32 missing as their measurements were below threshold of the BIS machine.

### Comparison of serum creatinine and cystatin C

The mean serum creatinine (scr) was 0.63 mg/dl (SD 0.15) with a range of 0.29 to 1.2mg/dl. The mean scr was significantly different according to sex, age, presence of stunting or viral suppression. The mean cystatin C was 0.81 mg/dl (SD 0.13) with a range of 0.51 to 1.39 mg/dl. The mean cystatin C was higher in males at 0.86 mg/dl versus 0.78 mg/dl in females but with no other differences (**Supplemental table 1**). Serum creatinine but not cystatin C was correlated with age and sex (**Figure 1**).

**Figure 1:**
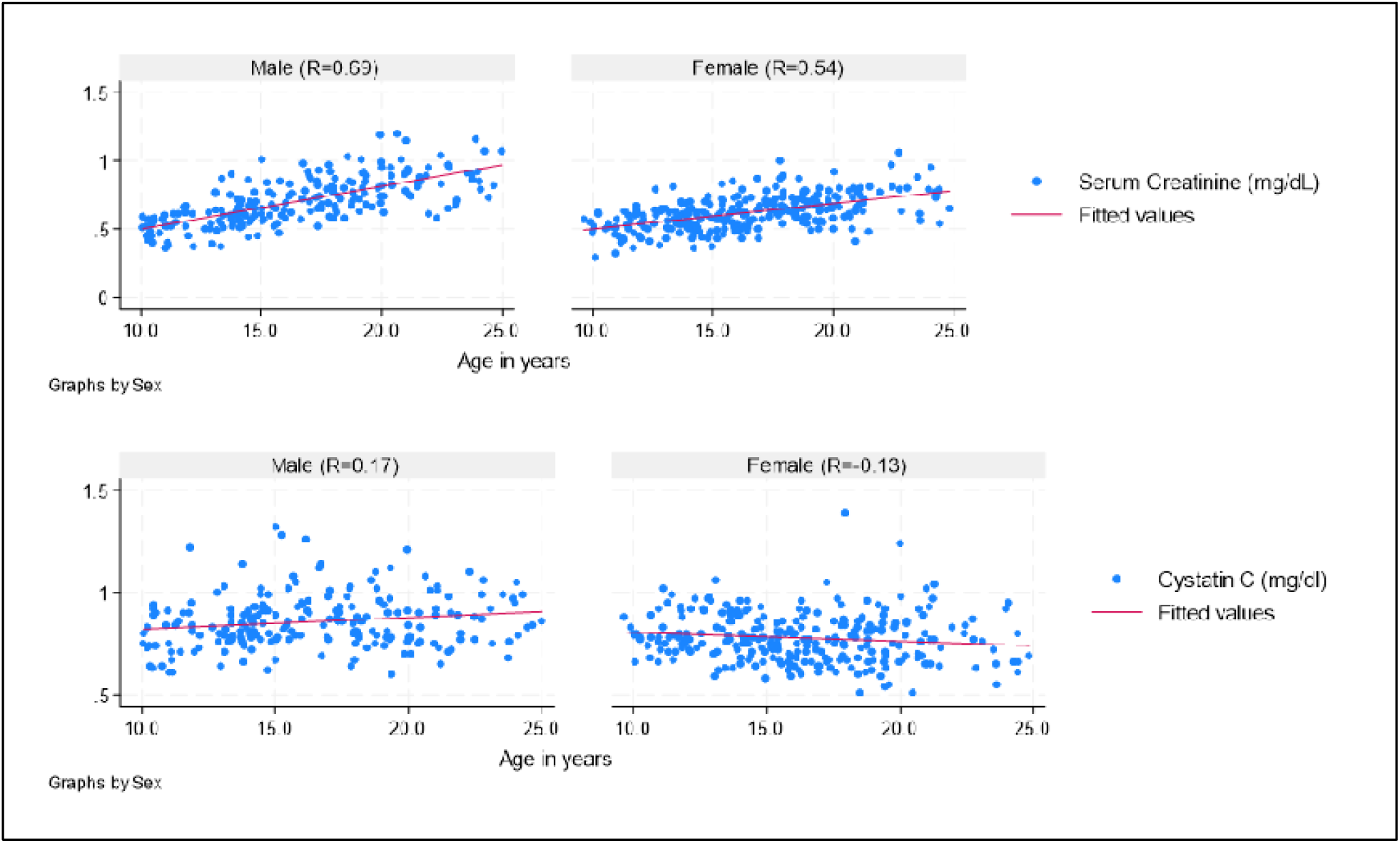
Relationship between serum creatinine and cystatin C and age for males and females

### Distribution of the eGFR

CKDEPI consistently gave higher eGFR readings for both creatinine and cystatin C, and the Schwartz cystatin C equation gave the lowest eGFR values (Figure 2).

**Figure 2.**
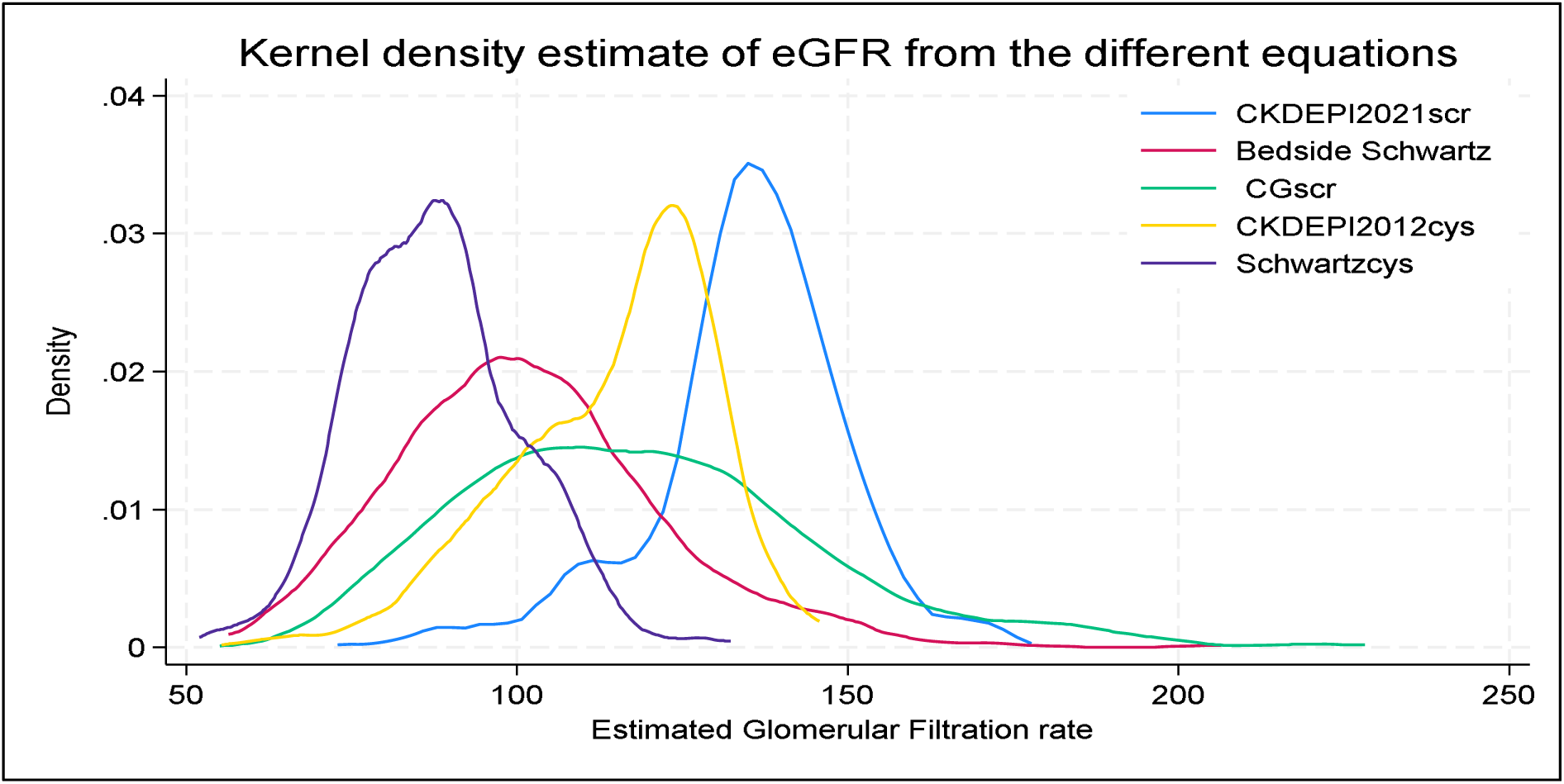
Kernel density plot showing the distribution of the eGFR according to different estimating equations and biomarkers.

### Prevalence of CKD using eGFR

CKD prevalence varied according to the eGFR cut-off, and the biomarker used. Using an eGFR<60ml/min/1.73m^2^ cut-off, the highest prevalence was with the Schwartz cystatin C equation (1.4%; 95% CI: 0.5-2.9% at baseline; 0.8%; 95% CI: 0.2-2.1% at 3-month follow-up) and the lowest with the CKDEPI 2021 equation (0%; 95% CI 0-0.07%). Similarly, using eGFR< 90ml/min/1.73m^2^ cut-off, the highest prevalence was with the Schwartz cystatin C equation (58.9%; 95% CI: 54.4-63.3%) and the lowest with CKDEPI (0.6%; 95% CI: 0.01-1.7%) (Figure 3). Prevalence using other eGFR equations ranged from 0% to 27.5% (**Supplementary Table 2**).

**Figure 3.**
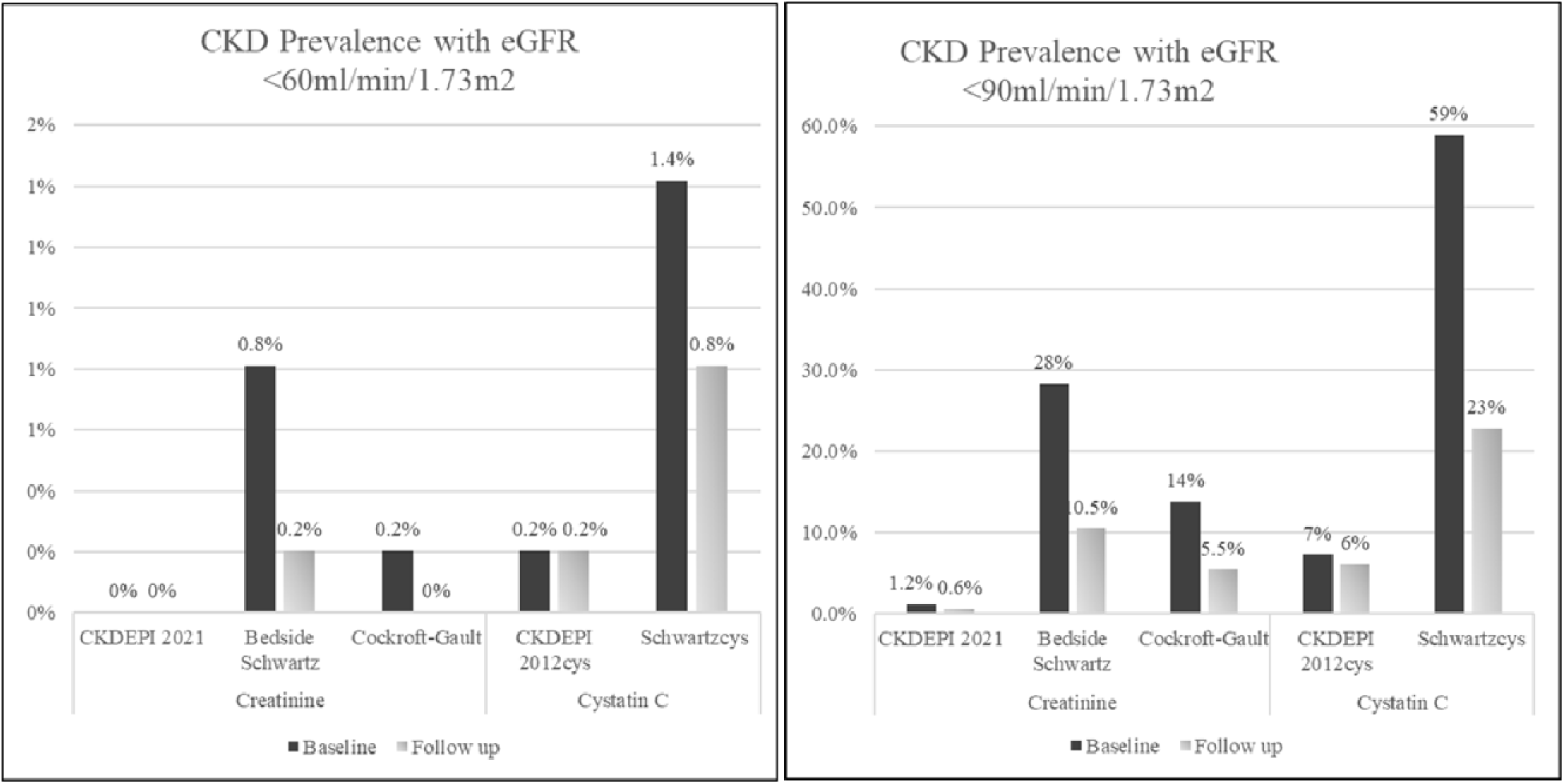
Prevalence of CKD according to the different estimating equations and biomarkers.

### Prevalence of CKD according to eGFR and ACR

All participants were staged according to combined baseline eGFR using cystatin C and ACR to assess risk of progression (28). Overall, 438 (88.5%) participants had low risk of progression (green), 53 (10.7%) had intermediate risk of progression (yellow) and 4 (0.8%) were at high risk of progression (orange) (**Table 2**).

**Table 2.**
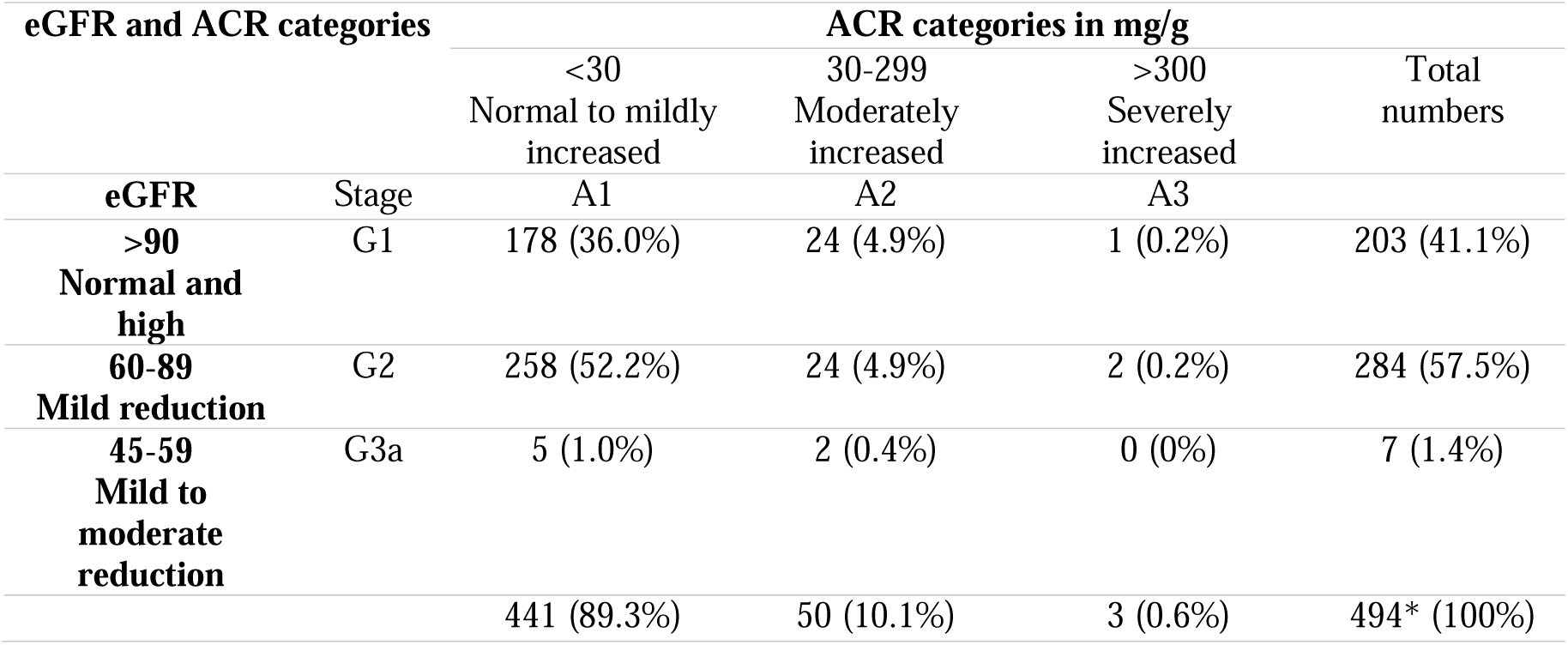
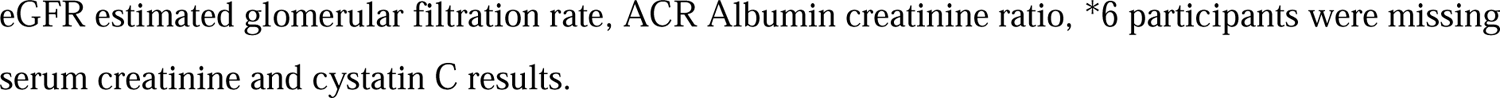
All participants’ CKD status staged according to estimated GFR from cystatin C and albumin creatinine ratio at baseline.

### Prevalence of CKD according to markers of kidney damage

Urinalysis showed that 143 (29%) participants had proteinuria on dipstick. Prevalence of proteinuria was similar for those with eGFR>90ml/min/1.73m^2^ and <90ml/min/1.73m^2^ (24.6% vs 31.7% p=0.17). At baseline, 10.1% (50) and at follow up, 3.8% (19) participants had an ACR >30mg/g.

### Factors associated with CKD

Factors associated with CKD varied with the equation and biomarker used for those with an eGFR <90ml/min/1.73m^2^ (**Table 3**) but was largely associated with male sex (with the exception of CKDEPI2021), viral non-suppression (by the cystatin C based equations), increasing age (by the CKDEPI and Bedside Schwartz equations), and being overweight (with the exception of the Cockroft-Gault equation). CKD was also associated with proteinuria (by the CKDEPI 2012 equation) and being on a TDF-based regimen (by the Bedside Schwartz equation). There was no evidence that CKD was associated with high blood pressure, muscle mass, and ACR. Results were similar when using CKD defined by eGFR<60ml/min/1.73m^2^ (**Supplementary Table 3**).

**Table 3.**
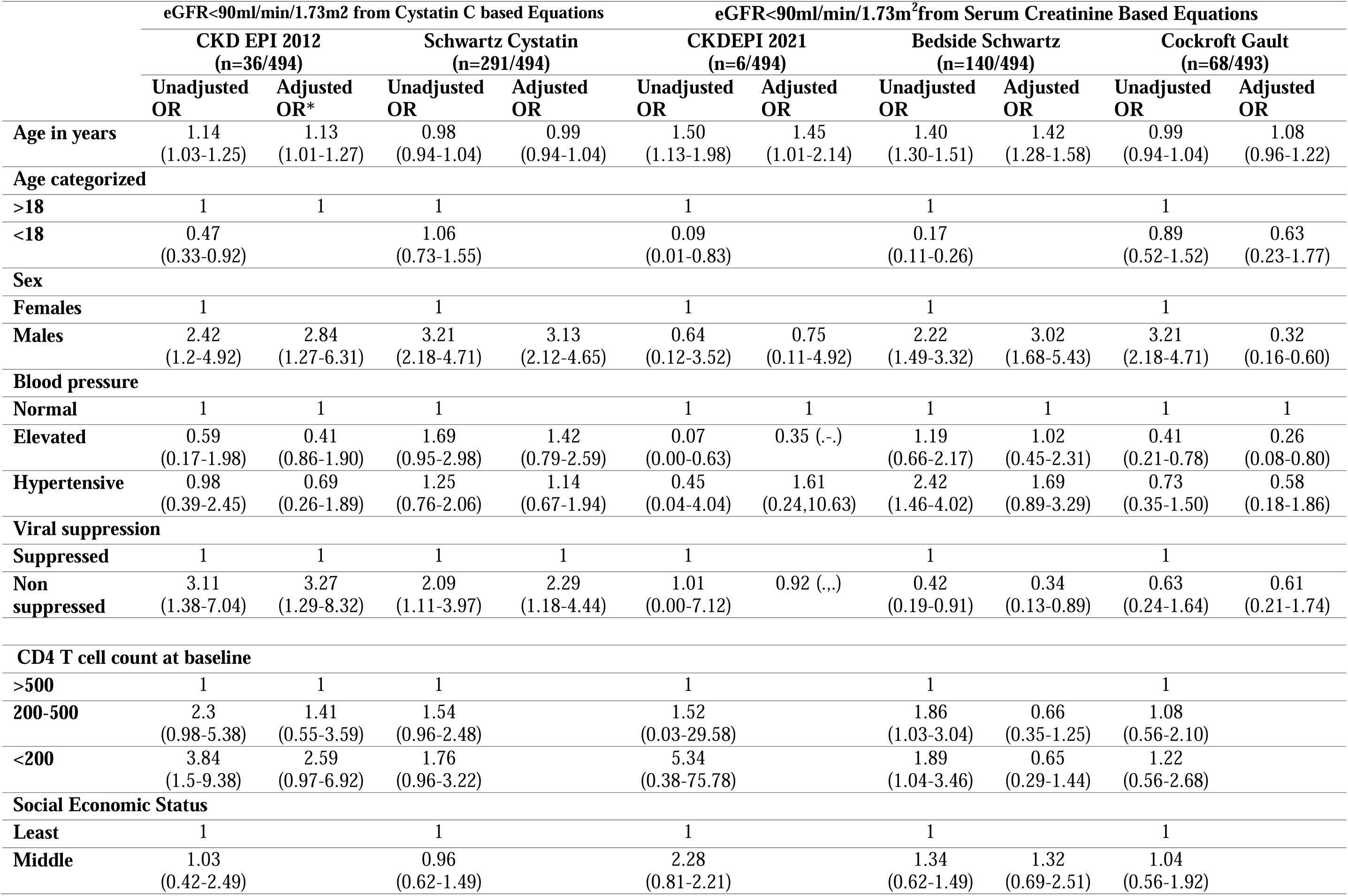
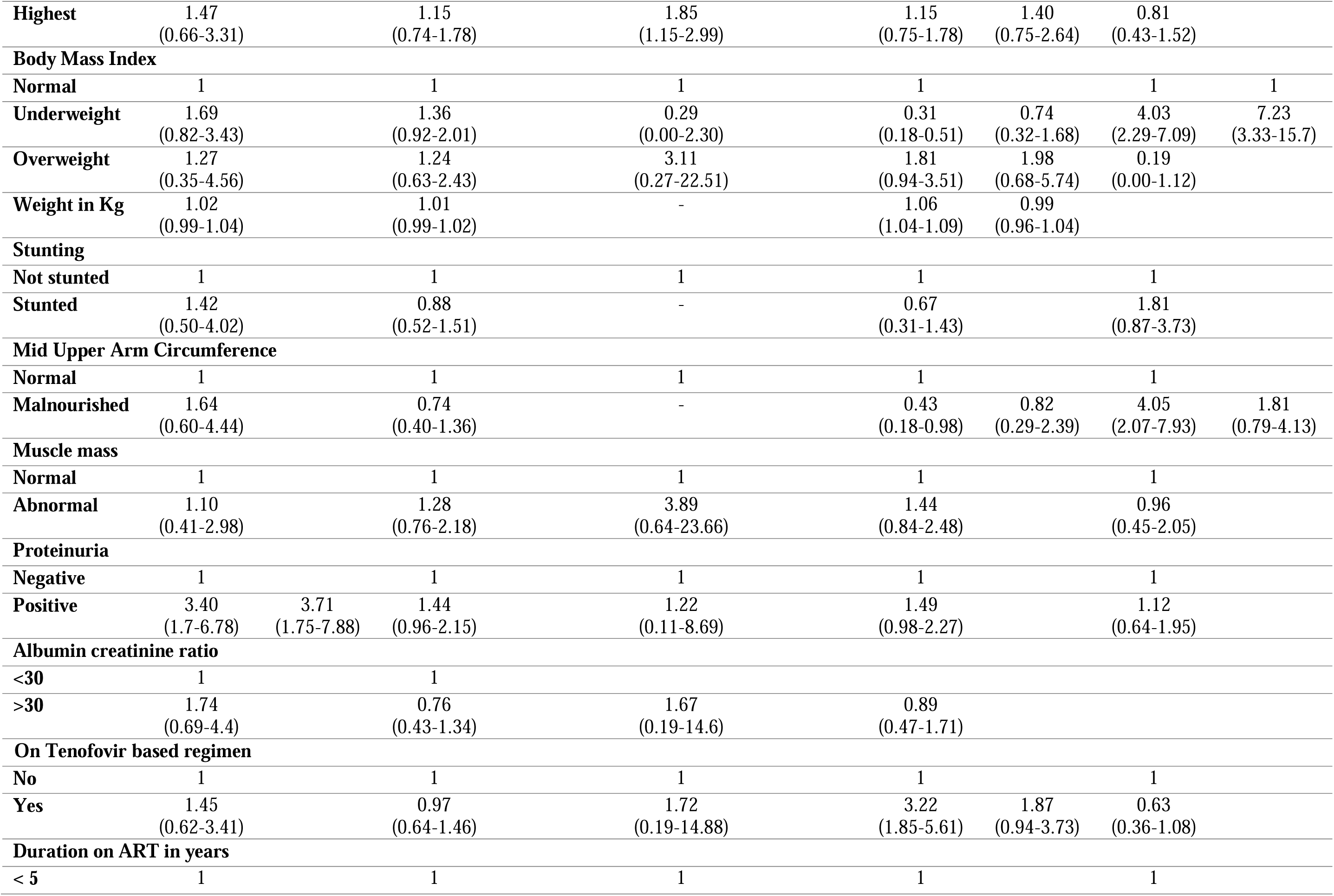
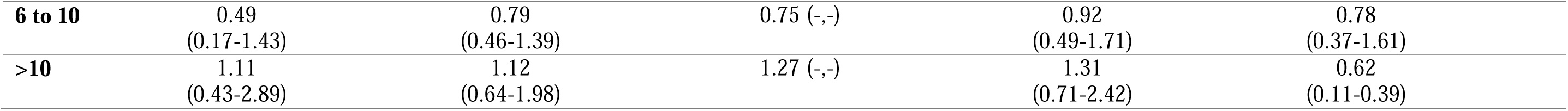
Factors associated with having CKD (eGFR<90**ml/min/1.73m^2^**) among study participants according to the different estimating equations and biomarkers. OR Odds ratio ART Anti-retroviral therapy. N Number, eGFR estimated glomerular filtration rate* Adjusted for age, sex, blood pressure, viral suppression proteinuria, baseline CD4 T cell count.

## Discussion

This is the first study to compare the prevalence and factors associated with CKD diagnosed by creatinine and cystatin C among YPLHIV in Uganda according to standard guidelines. We found highly variable prevalence depending on the definition, the estimating equation and the biomarker used. This was compounded by the commonly used GFR estimating equations being recommended for adults or children only, despite the highly variable physical and sexual maturity within this important age group where long-term disease management is critical. Using cystatin C eGFR measures consistently gave substantially higher prevalence of CKD: using the Schwartz cystatin equation approximately 60% of YPLHIV had eGFR <90mls/min/1.73m^2^. While dipstick proteinuria is anticipated in this population largely treated with anti-retroviral drugs, 10% of participants had substantially elevated levels of albuminuria. However, when participants with baseline abnormalities were remeasured at 3 months according to the gold-standard definition, overall prevalence of CKD was much lower.

The highest prevalence (59%) using an eGFR cutoff <90ml/min/1.73m^2^ at baseline which fell to 23% at three months follow-up, was very high. This is similar to a study done in 96 Nigerian YPLHIV aged 15 to 29 years which found 53.3% prevalence (36) and a Tanzanian study among 240 YPLHIV aged less than 14 years that showed a prevalence of 28% (37). When kidney function was determined by eGFR below 60ml/min/1.73m^2^ on two separate occasions at least three months apart, the prevalence of CKD was 0.8%. This is lower than in a study done in Zambia among children living with HIV aged 1 to 18 years that found a prevalence of 3.8% after 3 months (38). However, the children in this study were younger than in this study.

Using a eGFR cut off of less than 60ml/min/1.73m^2^ excludes a large proportion of YPLHIV who are already showing signs of impaired kidney function such as an ACR above 30mg/g, proteinuria and hypertension, and who would benefit from early intervention to halt progression of their kidney disease (39). Pottel et al. have shown that clinical manifestations of decreased kidney function in young people start at GFR less than 75ml/min/1.73m^2^; they recommend that the CKD definition should be revised to reflect this (40). KDIGO recommends that screening and surveillance for CKD be tailored to the specific high risk group (41). Our study suggests that using sequential estimation of eGFR over three months excludes YPLHIV at risk of CKD, and might be misleading to the public health response whose goal is to halt progression and to predict those who are in danger of kidney failure or development of cardiovascular complications (41). KDIGO further recommends that screening frequency should be based on the risk profile of the individual and potential to progress (42). YPLHIV have the potential to progress due to the continued insult to the kidney, one abnormal eGFR measurement that shows reduced kidney function should be sufficient for them to be followed up regularly and managed.

Estimating GFR in this population was challenging as the different estimating equations and biomarkers gave very different results. This was worse as one transitioned from equations meant for those below 18 years to those equations meant for adults above 18 years. The difference in the eGFR was wide even in the same individual. It is difficult to determine the true estimate for CKD among YPLHIV using these estimating equations yet knowing the true estimate is important to plan the public health response for CKD (15). Clinicians who seek to diagnose CKD and plan management may get confused about the true CKD status of an individual. Misdiagnosis and classification of YPLHIV removes the opportunity to intervene early to halt progression to kidney failure (43). However, it is not surprising that each of the estimating equations gave a different prevalence since each estimating equation reflects the characteristics of the population/dataset that was used to develop it (44). There is an urgent need to develop estimating equations for Africans living in Africa.

The use of GFR alone doesn’t predict progression or mortality risk and other markers of kidney damage such as albuminuria or proteinuria are used (13, 45). When ACR was used, the prevalence was 10.1%. This is lower than that reported among a Tanzanian cohort of YPLHIV aged 1 to 14 years which found a prevalence of 20.1% (37). However, the ACR was determined at a single time point and included younger children. Proteinuria prevalence was 29% which was high in such a young population. Proteinuria is an early marker of HIV associated nephropathy (46) and if persistent, is predictive of CKD status in children (47). However, we measured proteinuria only at baseline and yet two positive out of three readings are used to diagnose persistent proteinuria (41).

Cystatin C emerged as a better biomarker than serum creatinine as eGFR calculated from Cystatin C was above CKD stage 1 more consistently for all those that had an increased ACR, proteinuria or hypertension which are markers of abnormal kidney function (18). Cystatin C was recommended by a recent study in three countries (Uganda, Malawi, and South Africa) as the better biomarker in Africans (4). Cystatin C should be recommended for the diagnosis of CKD in YPLHIV as well.

We found that age, sex, and HIV viral non-suppression were associated with CKD and that proteinuria, CD4 cell count, blood pressure, and being on a TDF regimen were not associated. A study among perinatally infected YPLHIV in South Africa with a mean age of 12.0 years found sex, but not age or blood pressure were associated with CKD (48). Males were also found to have more CKD than females in a study in Zimbabwe (49). TDF use was also not associated with CKD status in a cohort of American children with CKD (50).

One of the strengths of this study is that we estimated the eGFR at two different time points more than three months apart as recommended by KDIGO and were able to ascertain those that actually had CKD according to the standard definition of CKD. However, most of the GFR estimating equations and normal serum creatinine have not been validated in YPLHIV in resource-limited settings especially in Africa and this makes it that much harder to determine the abnormal values in YPLHIV (4, 51). This could explain the low correlation between eGFR and the markers of kidney damage found in this study. We determined both markers of kidney damage (albuminuria and proteinuria) and function and could tell YPLHIV that were at risk of CKD progression. The biggest limitation is that we did not measure the GFR using either ioxehol or the nuclear tracers 99mTc-diethylenetriaminepentaacetic acid (DTPA) or ^51^Cr-EDTA (10) and so we are unable to tell how accurate the eGFR was.

## Conclusion

CKD prevalence among YPLHIV in Uganda varies widely depending on the biomarker and definition used. However, there is a substantial prevalence of albuminuria and reduced eGFR suggesting HIV programs should prioritize screening for CKD among YPLHIV. The definition of CKD and best biomarker to use in YPLHIV should be further investigated to optimise detection of those with early abnormalities of kidney function. Estimating equations should be validated against measured GFR in young people to define how best to estimate GFR across older children and young adults in Africa.

## List of abbreviations

ACR: Albumin Creatinine Ratio

AIDS: Acquired Immune Deficiency Syndrome

ALHIV: Adolescents living with HIV

ART: Anti-Retroviral Therapy

BIS: Bioimpedance Spectroscopy

BMI: Body Mass Index

CAKUT: Congenital abnormalities of the Kidney and Urinary Tract

CALHIV: Children and Adolescents living with HIV

CAP: College of American Pathologists

CBC: Complete Blood Count

CD4: Cluster of differentiation 4

CKD: Chronic Kidney Disease

CKD-EPI: Chronic Kidney Disease Epidemiology Collaboration

DM: Diabetes Mellitus

ESKD: End stage kidney disease

EKFC: European Kidney Function Consortium

FAS: Full Age Spectrum

GFR: Glomerular Filtration Rate

HB: Haemoglobin

HIV: Human Immunodeficiency Virus

HIVAN: HIV associated Nephropathy

HIVICK: HIV Immune Complex Kidney Disease

HW: Health Worker

KDIGO: Kidney Disease Improving Global Outcomes

KRT: Kidney replacement therapy

MDRD: Modification of Diet in Renal Disease

MOH: Ministry of Health

NCD’s: Non-Communicable Diseases

PCR: Protein Creatinine Ratio

PLHIV: People Living with HIV AIDS

RAAS: Renin Angiotensin Aldosterone Systems

RCT: Randomised Controlled Trials

SSA: Sub Saharan Africa

TB: Tuberculosis

UNAIDS: United Nations Joint AIDS program

USA: United States of America

WHO: World Health Organization

YPLHIV: Young People Living with HIV

## Declarations

### Consent for publication

Not applicable

### Availability of data and materials

The data supporting the findings of this study are openly available in repository https://datacompass.lshtm.ac.uk/.

### Competing interests

The authors declare no conflict of interest.

## Funding

Support for research was provided by Fogarty International Centre, National Institutes of Health (grant #2D43TW009771-06) HIV and co-infections in Uganda. HAW is funded by the UK Medical Research Council (MRC) and the UK Department for International Development (DFID) under the MRC/DFID Concordat agreement (Grant 1: MR/R010161/1). EN, Doctoral Research Fellow, NIHR131273 is funded by the NIHR for this research project. The views expressed in this publication are those of the authors and not necessarily those of the NIHR, NHS or the UK Department of Health and Social Care.

## Authors’ contributions

EN, LT, RK, CDC, DN, BC, YCM, HW contributed to the conceptualization and design of the study, data collection, analysis, and interpretation. EN, LT, YK drafted the manuscript. CDC, BC, RK, YCM edited the draft manuscript. HW was responsible for the overall supervision of this work. All authors reviewed and approved the manuscript before submission for publication.

## Data Availability

All data produced in the present study are available upon reasonable request to the authors

## Acknowledgements

The authors acknowledge the health workers and peers at the implementing facilities where this data was collected. The HIV program at the participating health facilities is supported by the President’s Emergency Plan for AIDS Relief (PEPFAR) through CDC.

## Notes

### Competing Interest Statement

The authors have declared no competing interest.

### Author Declarations

Ethical approval was received from the Uganda Virus Research Institute (UVRI) Research Ethics Committee (reference number GC/127/946), the Uganda National Council of Science and Technology (HS2578ES) and the London School of Hygiene and Tropical Medicine institutional review board (28797). Information about the study appropriate for adults, semi-literate adults and children was provided in an information booklet that was read to the participants and caregivers. All the participants more than 18 years of age provided a written informed consent. Those below 18 years of age provided assent and their caregivers provided written informed consent. If a child refused to provide assent even after their caregiver had provided consent, that child was not enrolled into the study. All participants had the option to withdraw at any point during the research. All participants with suspected CKD were referred to a nephrologist for management. Consent for publication: Not applicable

